# Emotion regulation in emerging adults with major depressive disorder and frequent cannabis use

**DOI:** 10.1101/2020.11.25.20238097

**Authors:** Emily S. Nichols, Jacob Penner, Kristen A. Ford, Michael Wammes, Richard W.J. Neufeld, Derek G.V. Mitchell, Steven G. Greening, Jean Théberge, Peter C. Williamson, Elizabeth A. Osuch

## Abstract

In people with mental health issues, approximately 20% have co-occurring substance use, often involving cannabis. Although emotion regulation can be affected both by major depressive disorder (MDD) and by cannabis use, the relationship among all three factors is unknown. In this study, we used fMRI to evaluate the effect that cannabis use and MDD have on brain activation during an emotion regulation task. Differences were assessed in 74 emerging adults aged 16-23 with and without MDD who either used or did not use cannabis. Severity of depressive symptoms, emotion regulation style, and age of cannabis use onset were also measured. Both MDD and cannabis use interacted with the emotion regulation task in the left temporal lobe, however the location of the interaction differed for each factor. Specifically, MDD showed an interaction with emotion regulation in the middle temporal gyrus, whereas cannabis use showed an interaction in the superior temporal gyrus. Emotion regulation style predicted activity in the right superior frontal gyrus, however, this did not interact with MDD or cannabis use. Severity of depressive symptoms interacted with the emotion regulation task in the left middle temporal gyrus. The results highlight the influence of cannabis use and MDD on emotion regulation processing, suggesting that both may have a broader impact on the brain than previously thought.

## 1. Introduction

Major depressive disorder (MDD) is a potentially debilitating psychiatric disorder with an estimated worldwide prevalence in emerging adults of 16-18% (1–3). Cannabis is the most commonly used recreational drug after alcohol and the highest prevalence of use is in teens and young adults (4). A recent study of Canadian middle-school age youth showed that cannabis use was strongly associated with internalizing mental health problems (viz., depression, anxiety) with an odds ratio of approximately 6.5 (5). There is some overlap in symptomatology between MDD and heavy cannabis use including anhedonia, changes in weight, sleep disturbance and psychomotor problems (6). A recent meta-analysis also found that adolescent cannabis use predicted depression and suicidal behaviour later in life (7). The link between mood disorders and cannabis use is complex, especially with respect to directionality; cannabis use is predictive of the onset of mood disorders in youth (8–11), even while some individuals use cannabis in an attempt to regulate the symptoms of depression (12, 13). The likelihood of developing MDD in heavy cannabis users who began at a young age has been estimated to be up to 8.3 times higher than in individuals who do not use cannabis (14). Emotion regulation, or the ability to modify one’s emotional experience to produce an appropriate response, has been shown to be maladaptive in teenagers and young adults with MDD and who use cannabis (15–18). For example, suppression is a maladaptive regulation style in which an individual inhibits expressing emotions, and is correlated with greater depressive symptoms in youth and adults (19). In contrast, reappraisal is an adaptive regulation style in which an individual changes their interpretation of a situation to alter the emotional impact, and is underutilized in emerging adults with MDD (17) and in those who are cannabis users (15).

In the context of MDD, studies have shown lower activity in brain areas involved in emotional processing when compared to healthy controls in the dorsolateral prefrontal cortex (dlPFC), ventrolateral prefrontal cortex (vlPFC), anterior cingulate cortex, as well as the basal ganglia (20–25). These findings fit well with models of emotion regulation and of MDD. Emotion regulation is thought to occur through a network of regions, beginning with affective arousal in the amygdala and basal ganglia, then projecting to frontal regions including the vlPFC and the insula, as well as other regions such as the superior temporal gyrus (STG) and angular gyrus (26). The vlPDC then begins the process of emotional appraisal, indicating the need for regulation to the dlPFC. From there, the dlPFC regulates the emotion and feeds forward to the angular gyrus, STG, and back to the amygdala and basal ganglia, all of which create a regulated emotional state (26–31). Disruption of the communication among these areas in individuals with MDD has been observed both in measures of resting state connectivity (32, 33) and in the suppression of activity within these frontal regions in association with over-activation of temporal regions such as the insula and hippocampus (24).

The prevalence of depressive symptoms in frequent cannabis users suggests that brain regions involved in emotion regulation may overlap with those affected by cannabis use. A study showing emotion regulation deficits in young, regular recreational cannabis users compared to non-users bolsters this hypothesis (15). Indeed, a meta-analysis showed that cannabis use was linked to brain activity abnormalities in the vlPFC, dlPFC, and dmPFC, orbital frontal cortex, ventral striatum, and thalamus (34). A recent review of the imaging literature indicated that adolescent cannabis users showed differences in frontal-parietal networks that mediate cognitive control (35). Further, emotion regulation deficits in frequent cannabis users were associated with abnormal neural activity in bilateral frontal networks as well as decreased amygdala-dorsolateral prefrontal cortex functional connectivity (15). Suppressed inferior frontal and medial PFC activation has been found in cannabis users during positive and negative emotional evaluation (36), as has suppressed activity levels in the amygdala (36, 37). The overlap in these brain regions, combined with weakened emotional regulation in people with both MDD and cannabis use, suggests that there may be an interaction between MDD and cannabis use on human brain function in the context of emotion regulation.

The aim of the present study was to examine the combined effect of MDD and cannabis use on the brain during emotion regulation in emerging adults, as well as how specific characteristics, such as degree of depressive symptoms and age of cannabis use onset, affect emotion processing. To address these questions, we employed an emotion regulation task while participants underwent functional magnetic resonance imaging (fMRI). We recruited individuals either with or without MDD, who either did or did not use cannabis frequently, and used a mixed effects approach to identify the unique contributions of each factor on emotion processing. Because both MDD and cannabis use have been shown to suppress activation within frontal regions during emotion regulation, we predicted that combined MDD and cannabis use would interact with emotion regulation within the vlPFC, dlPFC, and dmPFC, above and beyond the contribution of each factor alone. In contrast, we predicted that we would see a dissociation between MDD and cannabis use in temporal regions, with MDD showing increased activity levels and cannabis use showing suppression of activity during emotion processing. Finally, we predicted that severity of depressive symptoms, emotion regulation style, and age of cannabis use onset would each uniquely interact with emotion regulation, further elucidating the relationship between MDD, cannabis use, and the brain. 2.

## 2. Methods

### 2.1 Participants and questionnaires

Participants were recruited from the local community and through the First Episode Mood and Anxiety Program (FEMAP) in London, Ontario, Canada. The research ethics board at Western University, London, Ontario, Canada provided approval for the protocol. Written informed consent was obtained from participants after a complete description of the study was provided. Data were collected from 77 participants, with four participants removed from the analysis; three due to missing data and one due to an incidental finding, resulting in 73 participants aged 16-23 (*M* = 19.85, *SD* = 1.63; 39 female) for further analysis. Although our analyses here did not examine individuals by group, they can be summarized as 20 non-depressed, non/low cannabis-using controls, 20 patients with MDD, 20 non-depressed frequent cannabis users, and 17 frequent cannabis users with either active or recent MDD. Our previous studies used most of the same participants (38, 39). The treating psychiatrists made the psychiatric diagnoses, confirmed by the Structured Clinical Interview for Diagnosis, DSM-IV (Axis I, SCID-CV) (40). Cannabis use intensity has been stratified in numerous ways in previous research (41, 42); in the current study frequent use was defined as ≥ 4 times per week for at least 3 months preceding the study (38). Cannabis use was assessed by self-report and verified by urine screen to confirm all group assignments. Minimal lifetime cannabis use was allowed in the non-cannabis users because complete elimination would have been prohibitively restrictive in this demographic; non-significant use was defined as ≤ 3 times per month for the past year, though most of the non-users had even less frequent use (38). These limits were chosen to differentiate “experimentation” in controls from consistent cannabis use in the designated frequent cannabis users. In the current sample, only two “non-frequent users” had used cannabis in the past month; the first used it once, more than two weeks prior to the study. The second used it three times across a three-day period, more than three weeks prior to the study. Both participants tested negative for cannabis in their urine and indicated that they were not regular users.

Clinical information was gathered in-person by a member of the research team prior to fMRI data acquisition, as reported previously (38, 39). Relevant to the present study, the Emotion Regulation Questionnaire (ERQ) (19) was used to asses emotion regulation strategies and Hamilton Depression Rating Scale (HAM-D) (43) was used to assess severity of depression in all participants. Cannabis and alcohol quantities and age of onset of use were collected by administration of the Youth Risk Behavior Survey, 2009 version (44), which is self-report. Amongst individuals who used cannabis, there was no correlation between frequency of cannabis and alcohol use, measured by the number of days in the past month that they had used each substance (*r*(31) = −0.02, *p* = .899). Study eligibility included absence of head injury or serious medical illness (other than psychiatric diagnoses). Thirty-seven participants met the diagnostic criteria for a major depressive episode, with 32 experiencing a current episode and five participants having had one in the recent past (viz., within the last 12 months). Fifteen of these participants were currently on psychoactive medications, primarily selective serotonin reuptake inhibitors (SSRIs), all of whom had current MDD. Medication dose was stable for three weeks before fMRI data acquisition. None of the remaining 40 participants met criteria for a current or past depressive episode.

### 2.2 Emotion regulation paradigm

The emotion regulation fMRI task, adopted from Greening et al. (25), was designed to have participants actively alter their feelings elicited by sad (negative) and happy (positive) emotional scenes. Twenty negative and 20 positive emotional scenes were taken from the International Affective Picture System (45) for this study. The task involved viewing both negative and positive emotional scenes while being instructed to either simply view the scene (attend) or actively alter their feelings while viewing the scene (reduce negative feelings during negative scenes and enhance positive feelings during positive scenes). The four task conditions were therefore attend-negative, reduce-negative, attend-positive, and enhance-positive.

During the reduce-negative task condition participants were instructed to ‘acknowledge that the scene is negative. However, it does not affect you, things do not stay this bad, and the scene does not reflect the whole world’ and during the enhance-positive task condition participants were instructed to ‘acknowledge that the scene is positive. Further, that it does affect you, things can and do get even better and the scene does reflect the real world’ (25). This paradigm attempts to target and modify the negative thought tendencies about self, the world, and the future that are typical for depressed patients (46).

Participants were trained and practiced the paradigm before being scanned. During 4 imaging runs each participant completed 20 trials of each task condition (80 trials total). The 20 negative and 20 positive emotional scenes were displayed twice, once during the attend condition and again during the regulate condition. Participants never saw the same picture twice in the same run. To help mitigate any order affects, the trial order in each run was set as 4 independent runs and these were counterbalanced across subjects.

### 2.3 Imaging data acquisition

All magnetic resonance imaging (MRI) scans were acquired using the Lawson Health Research Institute’s 3T MRI scanner (Siemens Verio, Erlangen, Germany) with a 32-channel head coil. T1-weighted anatomical images were acquired covering whole brain with 1 mm isotropic resolution; anatomical images were used to orient the functional MRI (fMRI) images 6° coronal to the AC–PC plane and as a reference for spatial normalization. Blood oxygen level dependent (BOLD) activation was measured using fMRI images acquired with a 2D multi-slice, gradient-echo, echo-planar T2*-weighted scan (TR=2s, TE=20 ms, flip angle = 90°, FOV = 256×256×144 mm3, 4 mm isotropic resolution); 4 runs of 200 functional volumes totaled approximately 26 minutes for the scan.

### 2.4 Data preprocessing and analysis

Results included in this manuscript come from preprocessing performed using *fMRIPrep* 1.3.2 (RRID:SCR_016216) (47, 48), which is based on Nipype 1.1.9 (RRID:SCR_002502) (49, 50). The *fMRIPrep* pipeline uses a combination of tools from well-known software packages, including <u>FSL</u>, <u>ANTs</u>, <u>FreeSurfer</u> and <u>AFNI</u>. This pipeline was designed to provide the best software implementation for each state of preprocessing (47, 48).

#### 2.4.1 Anatomical data preprocessing

T1-weighted (T1w) images were corrected for intensity non-uniformity (INU) with N4BiasFieldCorrection (51), distributed with ANTs 2.2.0 (52) (RRID:SCR_004757). The T1w-reference was then skull-stripped with a Nipype implementation of the antsBrainExtraction.sh workflow (from ANTs), using OASIS30ANTs as target template. A T1w-reference map was computed after registration of 2 T1w images (after INU-correction) using mri_robust_template (FreeSurfer 6.0.1) (53). Brain surfaces were reconstructed using recon-all (FreeSurfer 6.0.1, RRID:SCR_001847) (54), and the brain mask estimated previously was refined with a custom variation of the method to reconcile ANTs-derived and FreeSurfer-derived segmentations of the cortical gray-matter of Mindboggle (RRID:SCR_002438) (55). Spatial normalization to the ICBM 152 Nonlinear Asymmetrical template version 2009c (RRID:SCR_008796) (46) was performed through nonlinear registration with antsRegistration (ANTs 2.2.0), using brain-extracted versions of both T1w volume and template. Brain tissue segmentation of cerebrospinal fluid (CSF), white-matter (WM) and gray-matter (GM) was performed on the brain-extracted T1w using fast (FSL 5.0.9, RRID:SCR_002823) (57).

#### 2.4.2 Functional data preprocessing

The functional data were also preprocessed according to the *fMRIPrep* pipeline. For each of the BOLD runs per subject, the following preprocessing was performed. First, a reference volume and its skull-stripped version were generated using a custom methodology of *fMRIPrep*. The BOLD reference was then co-registered to the T1w reference using bbregister (FreeSurfer) which implements boundary-based registration (58). Co-registration was configured with nine degrees of freedom to account for distortions remaining in the BOLD reference. Head-motion parameters with respect to the BOLD reference (transformation matrices, and six corresponding rotation and translation parameters) are estimated before any spatiotemporal filtering using mcflirt (FSL 5.0.9) (59). BOLD runs were slice-time corrected using 3dTshift from AFNI 20160207 (60) (RRID:SCR_005927). The BOLD time-series, were resampled to surfaces on the following spaces: fsaverage5. The BOLD time-series (including slice-timing correction when applied) were resampled onto their original, native space by applying a single, composite transform to correct for head-motion and susceptibility distortions. These resampled BOLD time-series will be referred to as preprocessed BOLD in original space, or just preprocessed BOLD. The BOLD time-series were resampled to MNI152NLin2009cAsym standard space, generating a preprocessed BOLD run in MNI152NLin2009cAsym space. First, a reference volume and its skull-stripped version were generated using a custom methodology of fMRIPrep. Several confounding time-series were calculated based on the preprocessed BOLD: framewise displacement (FD), DVARS and three region-wise global signals. FD and DVARS are calculated for each functional run, both using their implementations in Nipype (following the definitions by (61)). The three global signals are extracted within the CSF, the WM, and the whole-brain masks. Additionally, a set of physiological regressors were extracted to allow for component-based noise correction (CompCor) (62). Principal components are estimated after high-pass filtering the preprocessed BOLD time-series (using a discrete cosine filter with 128s cut-off) for the two CompCor variants: temporal (tCompCor) and anatomical (aCompCor). Six tCompCor components are then calculated from the top 5% variable voxels within a mask covering the subcortical regions. This subcortical mask is obtained by heavily eroding the brain mask, which ensures it does not include cortical GM regions. For aCompCor, six components are calculated within the intersection of the aforementioned mask and the union of CSF and WM masks calculated in T1w space, after their projection to the native space of each functional run (using the inverse BOLD-to-T1w transformation). The head-motion estimates calculated in the correction step were also placed within the corresponding confounds file. All resampling can be performed with a single interpolation step by composing all the pertinent transformations (i.e. head-motion transform matrices, susceptibility distortion correction when available, and co-registrations to anatomical and template spaces). Gridded (volumetric) resampling was performed using antsApplyTransforms (ANTs), configured with Lanczos interpolation to minimize the smoothing effects of other kernels (63). Non-gridded (surface) resampling was performed using mri_vol2surf (FreeSurfer).

Many internal operations of fMRIPrep use Nilearn 0.5.0 (RRID:SCR_001362) (64), mostly within the functional processing workflow. For more details of the pipeline, see the section corresponding to workflows in fMRIPrep’s documentation.

#### 2.4.3 Statistical analysis

Data analysis was conducted in AFNI Version AFNI_20.0.18 ‘Galba’ (60, 65, 66). The first level general linear model was conducted via *3dDeconvolve* to generate contrast maps for each individual participant, including a regressor-of-interest for each of the 4 task conditions (attend-negative, reduce-negative, attend-positive, enhance-positive). Six motion parameters (three rotation, three translation) were included as regressors of no-interest, as were the six aCompCor parameters. All regressors were produced by convolving a hemodynamic response function with a standard boxcar design. This generated beta-weight values at each voxel location for each of the four task conditions to carry forward to group analysis (2nd-level). Following first-level analysis, data were smoothed using a 6 mm gaussian kernel (AFNI *3dBlurToFWHM*), for a final average smoothing level of 8.18 mm.

For each of the following analyses, a whole-brain mask excluding the cerebellum was used. All analyses were performed using the AFNI function *3dLME* (67), a group analysis program that performs linear mixed effects (LME) analysis on data with multiple measurements per participant. The primary analysis tested the effects of cannabis use and MDD diagnosis on emotion regulation. The model was specified as follows: task condition (attend-negative, reduce-negative, attend-positive, enhance-positive), cannabis use (frequent/low or none), MDD diagnosis (yes/no), including two- and three-way interaction terms, were included as variables of interest. Medication use (yes/no), age, and number of alcoholic drinks consumed in the last 28 days as regressors. Sex was not included as a regressor due to high collinearity with cannabis use. Numeric variables (i.e., age and alcohol use) in this analysis and all subsequent analyses were mean-centered. A random effect of participant was included in the model, and a marginal sum of squares was used.

Three secondary analyses were then conducted. First, we examined the interaction between emotion regulation style and task-condition in the full sample. Similar to the main analysis, an LME model was specified with a condition x ERQ score interaction term, and age, alcohol, and medication use included as regressors. The ERQ score involved subtracting the maladaptive emotional style (suppression subscale score) from the adaptive style (reappraisal sub scale score). Thus, higher ERQ scores indicated more adaptive emotion regulation than lower scores. Two participants were excluded from this analysis due to missing ERQ score data.

Next, we examined the relationship between HAM-D score and BOLD-signal activation during the emotion regulation task. Here, only individuals with an active MDD diagnosis were included (n = 28). The LME model was specified with a condition x HAM-D score interaction, and age, alcohol, and medication were included as regressors.

Finally, the effects of early-onset cannabis use on task-related BOLD signal activation were examined. Here, we only included individuals who actively used cannabis (n = 34). We tested our hypothesis that early-onset cannabis use would have pronounced negative effects by grouping subjects into early-onset (under 15 years of age, n = 12) versus late onset (over 15 years of age, n = 22). LME analysis is well-suited for such unbalanced groups (68–70). We then identified where early-onset cannabis users had greater or lower activation than late-onset users. The LME model was specified with a condition x age of onset interaction, and age, alcohol, and medication were included as regressors.

For second-level analyses, the minimum cluster-size threshold was determined in two steps. First, we estimated the smoothness of the residuals for each subject output by *3dDeconvolve* using the autocorrelation function (ACF) option (AFNI *3dFWHMx*), and the mean smoothness level was calculated. Next, minimum cluster size was determined using a 10,000 iteration Monte Carlo simulation (AFNI *3dClustSim*) at a voxelwise alpha level of *p* = 0.05. Correction for multiple comparisons at *p* = 0.05 was achieved by setting a minimum cluster size of 64 voxels. Post-hoc contrasts were FDR corrected.

## 3. Results

### 3.1 Linear Mixed Effects – Cannabis Use, MDD, and Emotion Regulation

We first identified regions that showed activity modulated by cannabis use, MDD, and task condition. As reported in Table 1 and Figure 1A, there was a main effect of MDD in the left supramarginal gyrus, with individuals with MDD showing significantly greater activation than those without MDD (*t*(51.92) = −3.07, *p* = .003). As shown in Figure 2, there was also a main effect of condition in the left inferior parietal lobe, left middle frontal gyrus, right insula (negative reduce greater than rest), and left inferior frontal gyrus, with the direction of each effect shown in Figure 2B-H.

**Table 1.**
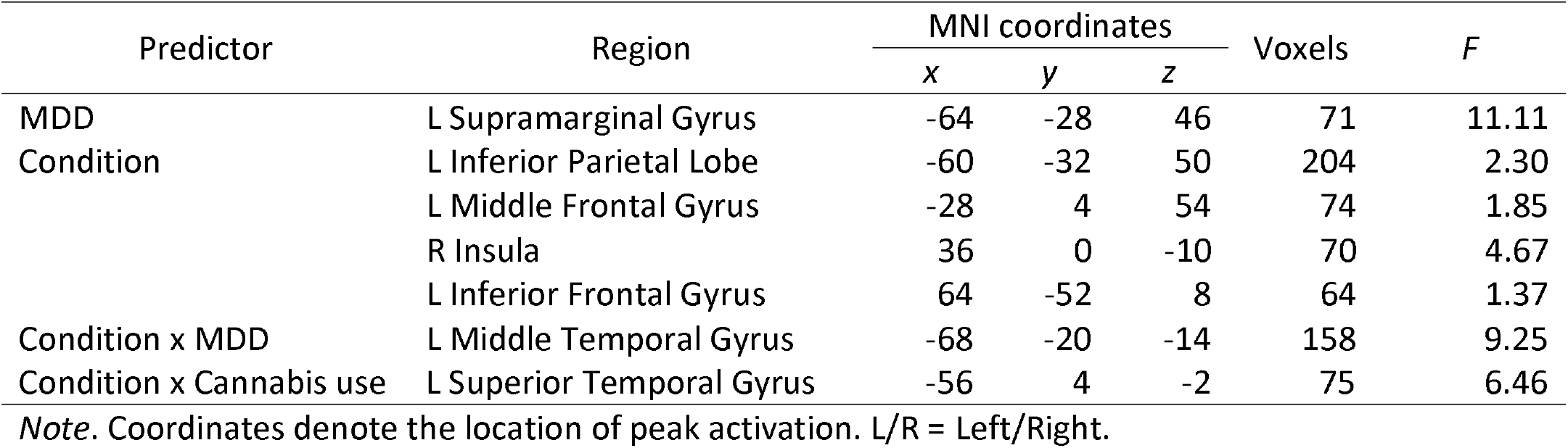
Clusters of significant activation

**Figure 1.**
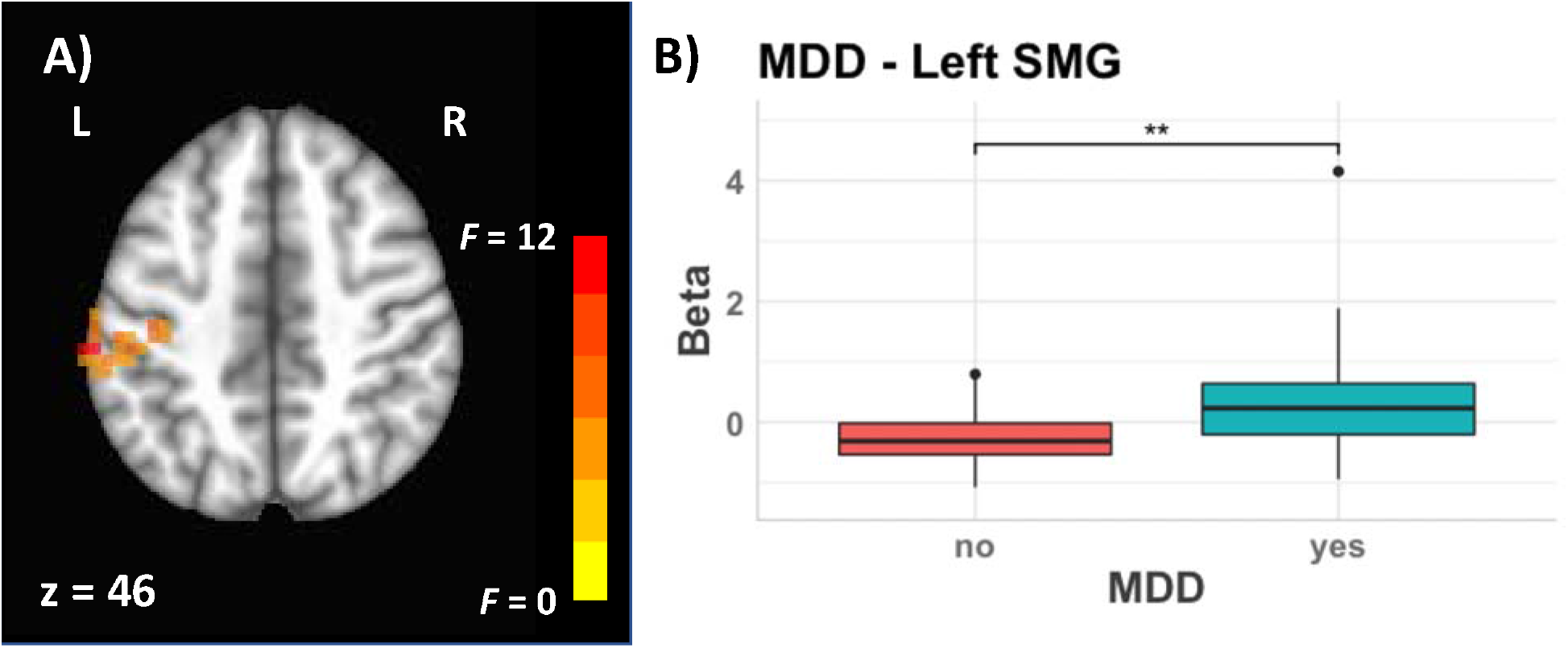
The effect of MDD on the brain. Statistical maps are thresholded at *p* = .05, overlaid on an MNI brain atlas. L = Left. Cluster locations and sizes are reported in Table 1. Boxplot shows betas in single voxel with peak activation. ** *p* ≤ .01. Medians are depicted as thick black horizontal lines within the boxes. 1^st^ and 3^rd^ quartiles are depicted as the lower and upper edges of the box, respectively. Lower and upper whiskers extend to the smallest and largest value within 1.5 * IQR, respectively. Outlying values beyond these ranges are plotted individually.

**Figure 2.**
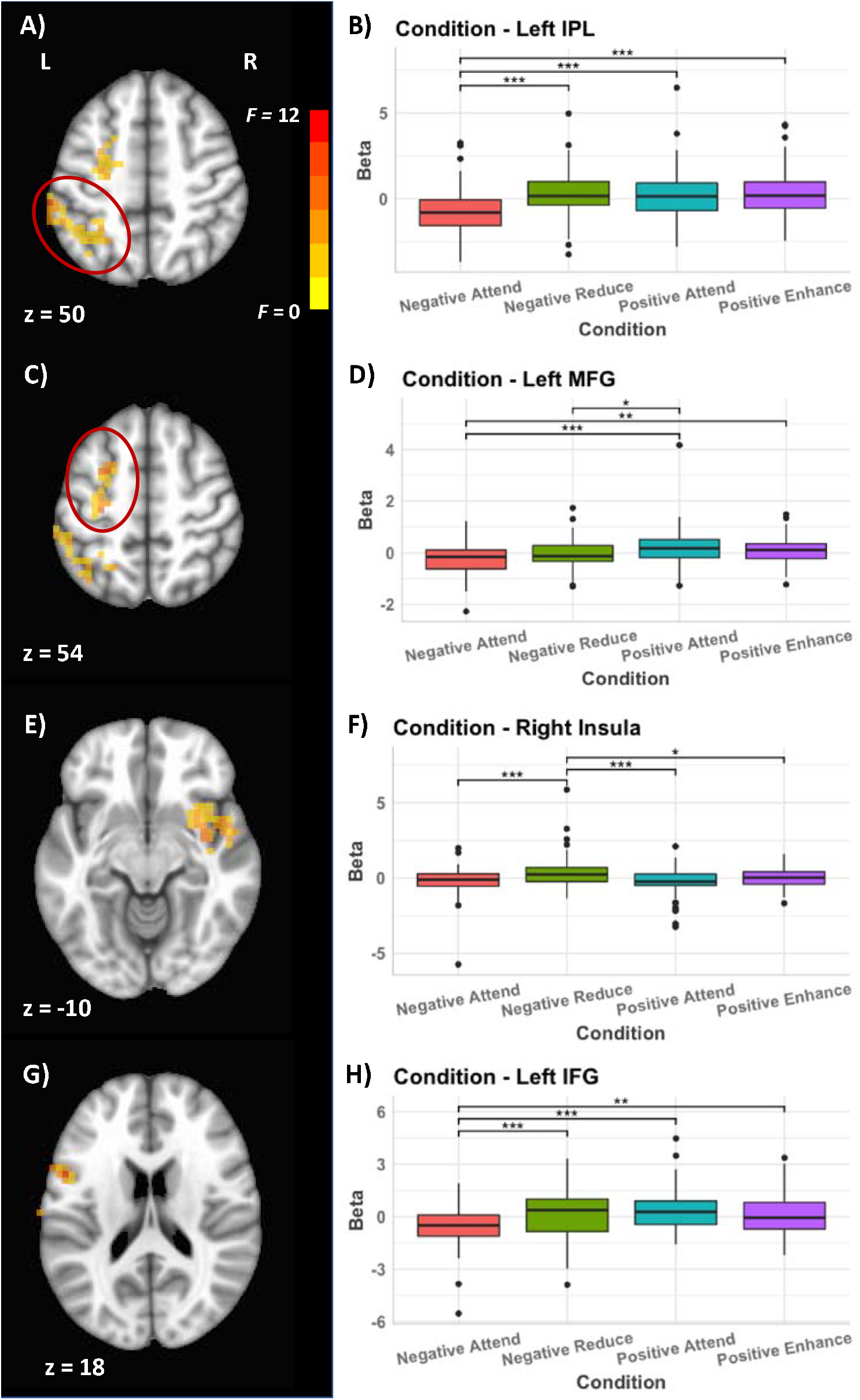
The effect of emotion regulation task condition on the brain. Brain maps and contrasts are given for the left inferior parietal lobe (IPL) in A) and B), the left middle frontal gyrus (MFG) in C) and D), the right insula in E) and F), and the left inferior frontal gyrus (IFG) in G) and H). Statistical maps are thresholded at *p* = .05, overlaid on an MNI brain atlas and show F-values. L = Left. Cluster locations and sizes are reported in Table 1. Boxplots show betas in single voxel with peak activation. * *p* ≤ .05, ** *p* ≤ .01, *** *p* ≤ .001. Medians are depicted as thick black horizontal lines within the boxes. 1^st^ and 3^rd^ quartiles are depicted as the lower and upper edges of the box, respectively. Lower and upper whiskers extend to the smallest and largest value within 1.5 * IQR, respectively. Outlying values beyond these ranges are plotted individually.

When examining interaction effects, there was a significant condition x MDD interaction in the left middle temporal gyrus (MTG). As can be seen in Figure 3B, all conditions showed increased activity in individuals with MDD, except for the positive attend condition in which they showed decreased activity. We also found a significant condition x cannabis use interaction in the left superior temporal gyrus (STG), shown in Figure 3C. As can be seen in Figure 3D, while the two emotionally positive conditions led to greater activity in individuals who use cannabis, the opposite was true for the emotionally negative conditions, with individuals who use cannabis showing lower activity. There was no significant 3-way interaction, no cannabis x MDD interaction, and no main effect of cannabis use.

**Figure 3.**
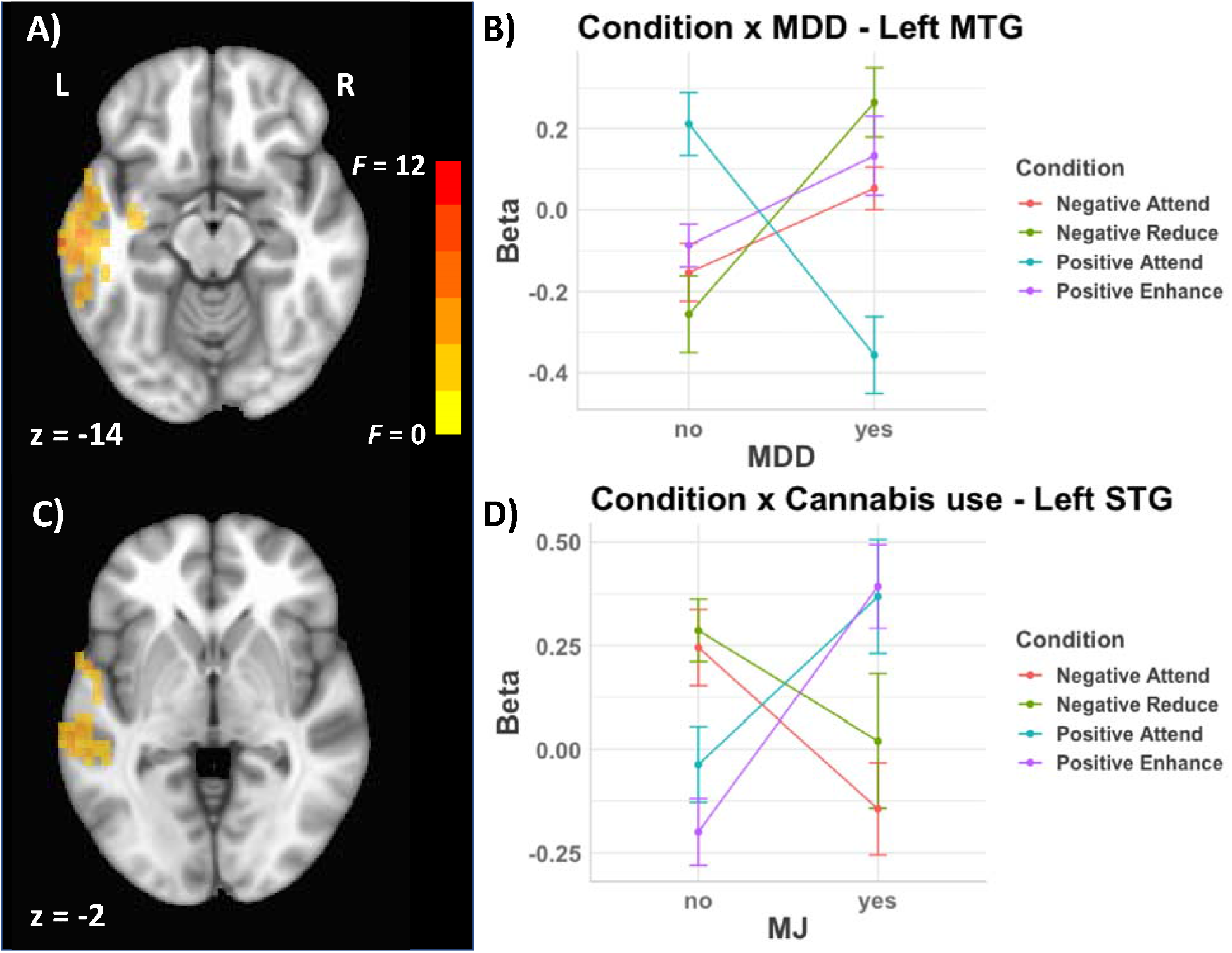
The relationship between MDD, cannabis use, and emotion regulation. The significant interaction between task condition and MDD is shown in A) and B), and the significant interaction between task condition and cannabis use is shown in C) and D). L = Left, statistical maps show *F-values*. Cluster locations and sizes are reported in Table 1.

### 3.4 Linear Mixed Effects – ERQ Score and Emotion Regulation

We examined emotion ratings during scanning using a linear mixed effects analysis with depression (yes/no), cannabis use (frequent/low or none), and trial condition as factors in the model, as well as all interaction terms. There was no main effect of depression on emotion ratings (*F*(1,67) = 2.18, *p* = 0.144), nor of cannabis use (*F*(1,67) = 0.14, *p* = .709), and no significant interactions. We next examined ERQ scores using a 2-factor ANOVA with depression and cannabis use as factors, as well as the interaction term. ERQ scores showed a significant main effect of depression (*F*(1,67) = 21.50, *p* < .001), no main effect of cannabis use, and no significant interaction. Post-hoc t-tests indicated that individuals with MDD had lower scores than individuals without MDD (*t*(69) = −4.72, *p* < .001).

When examining how ERQ score and task condition predicted brain activity in the full sample, we found a main effect of ERQ score in the left calcarine sulcus and right SFG, shown in Figure 4A. As can be seen in Figure 4B and 4C, both regions showed a decrease in activation with increasing ERQ score. Similar to the main analysis, we also found a main effect of condition in the left inferior parietal lobe and left inferior frontal gyrus. In both regions, the negative attend condition showed significantly lower activity than the other conditions (all *p*’s < .001). There was no significant interaction between ERQ score and task condition.

**Figure 4.**
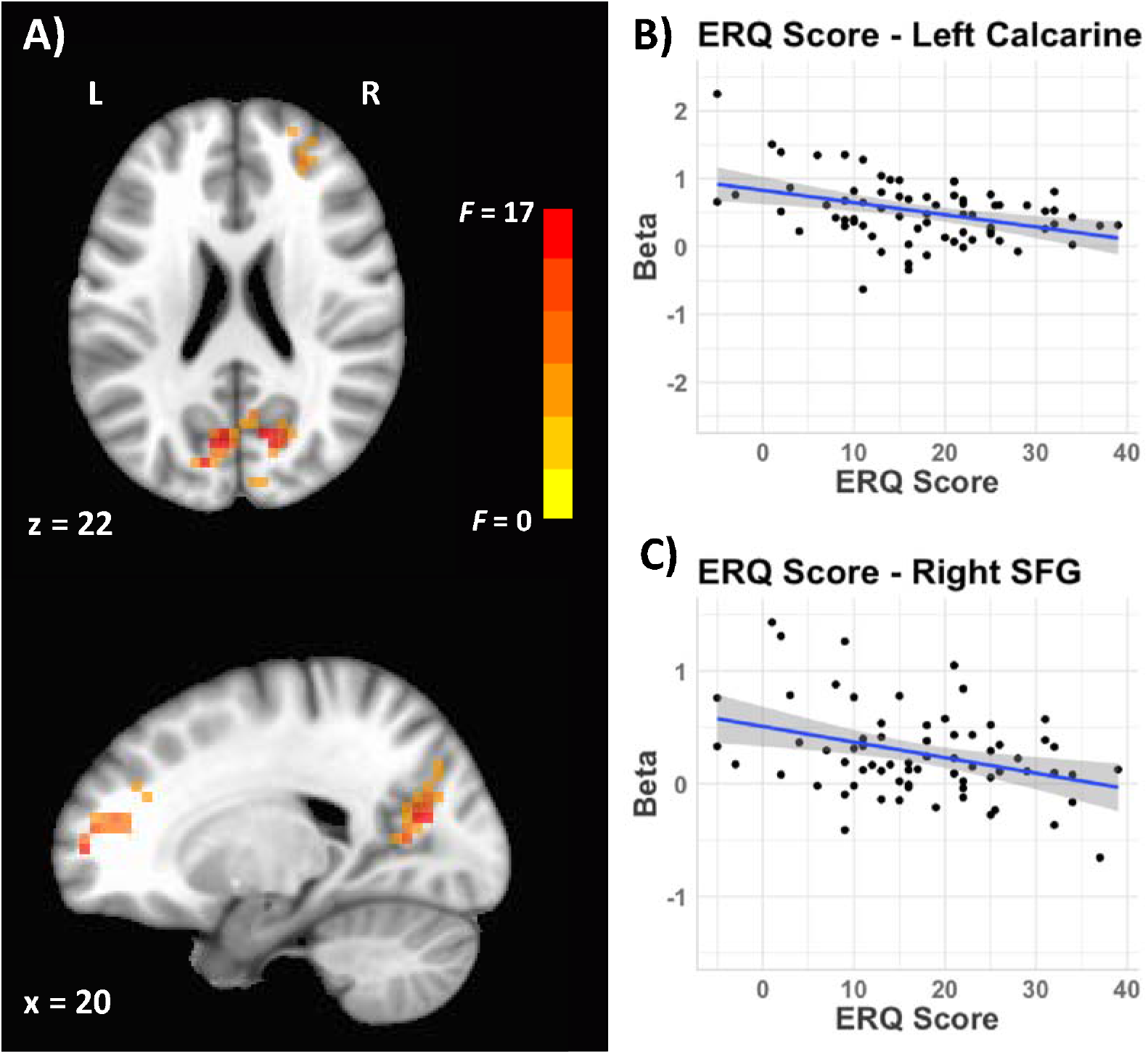
The relationship between emotion regulation style and brain activity during an emotion regulation task. A) Shows the activity significantly predicted by ERQ score in the left Calcarine sulcus and the right superior frontal gyrus. B) and C) show the scatterplots of beta values and ERQ score within each region. Statistical maps are thresholded at *p* = .05, overlaid on an MNI brain atlas. L = Left. Cluster locations and sizes are reported in Table 2.

### 3.5 Linear Mixed Effects – HAM-D Score and Emotion Regulation

Next, we examined how severity of MDD and task condition predicted brain activity in individuals with current MDD (n = 28). As shown in Figure 5, we found a significant condition x HAM-D score interaction in the left middle temporal gyrus, driven by an increase in activity in the negative reduce condition with increasing HAM-D score. There was also a significant main effect of HAM-D score in the left temporal pole, although this cluster overlapped with the interaction. In this reduced sample, there was also a significant effect of condition in the right MTG, left postcentral gyrus, and left temporal pole.

**Table 2.**
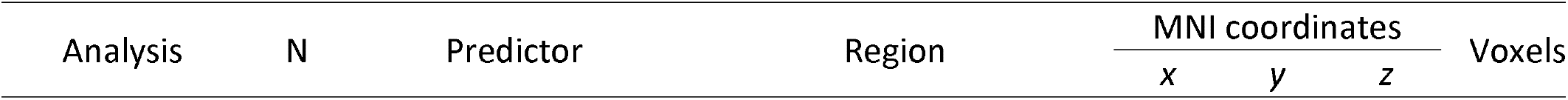
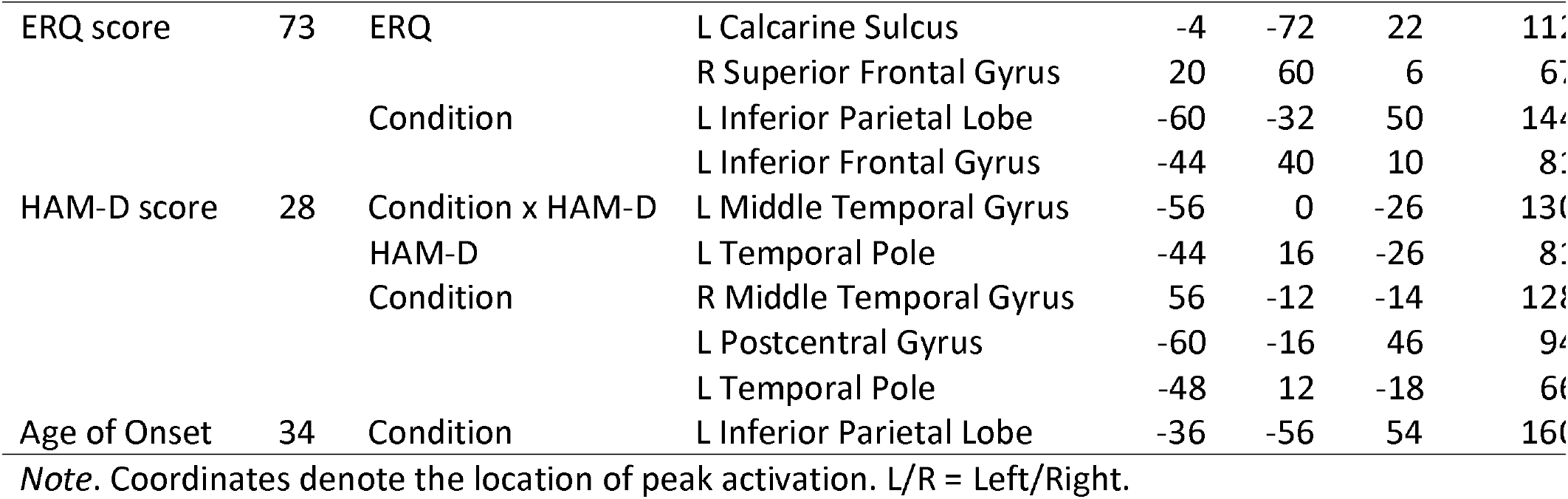
Clusters of significant activation in the secondary analyses of ERQ score, HAM-D score, and age of cannabis use onset in frequent cannabis users

**Figure 5.**
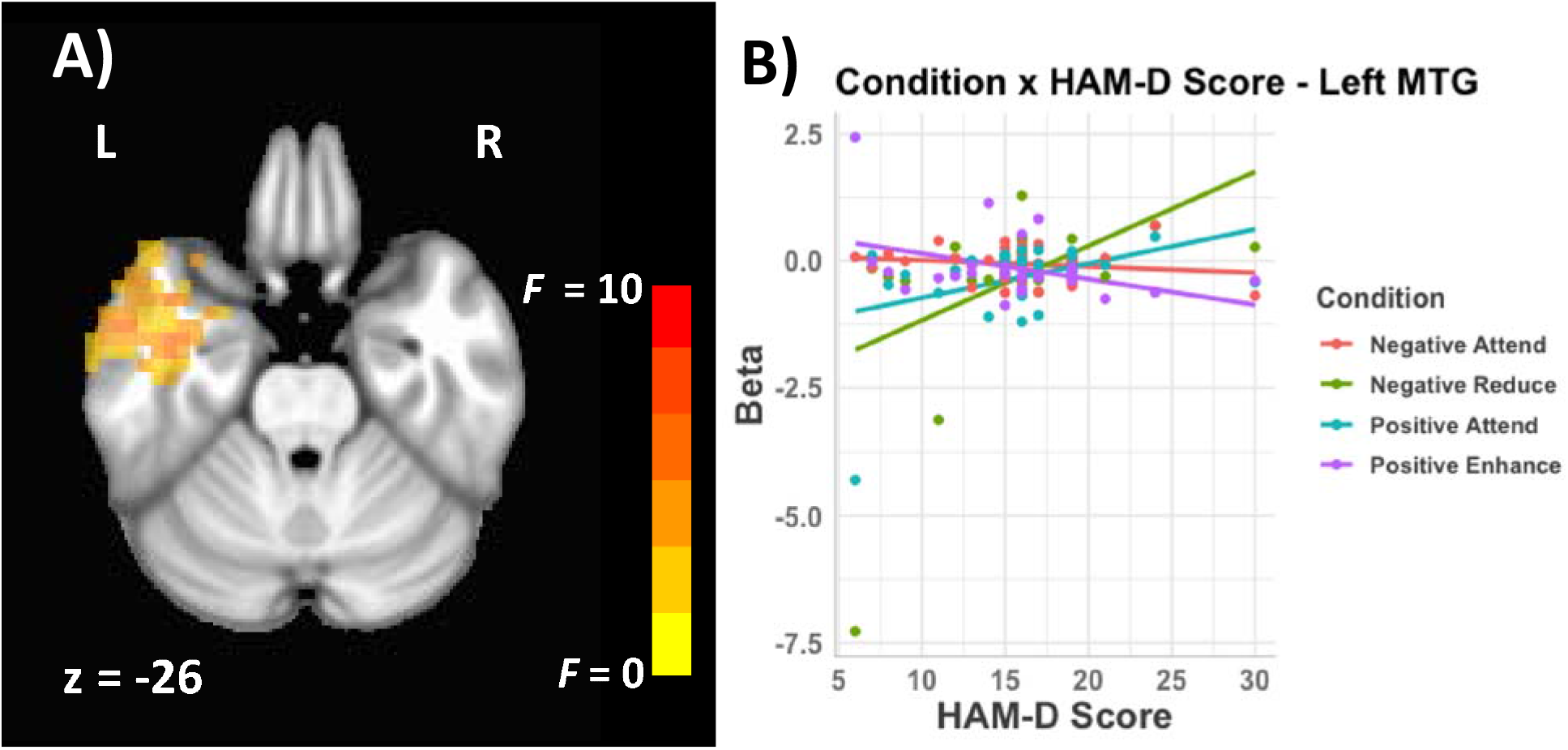
The relationship between HAM-D score and emotion regulation. Statistical maps are thresholded at *p* = .05, overlaid on an MNI brain atlas. L = Left. Cluster locations and sizes are reported in Table 2.

### 3.6 Linear Mixed Effects – Age of Cannabis Use Onset and Emotion Regulation

Finally, we examined how age of cannabis use onset and task condition predicted brain activity in individuals who were frequent users of cannabis (N = 34). Only task condition showed a significant effect in the left inferior parietal lobe. There was no significant age of onset x condition interaction, and no main effect of age of onset.

## 4. Discussion

The current study used an fMRI paradigm of positively- and negatively-valenced emotional scenes to investigate the individual and combined effects of MDD and frequent cannabis use on emotion regulation. We also conducted several secondary analyses to explore how the various characteristics of emotion regulation, MDD, cannabis use and age of onset of cannabis use further contribute to emotion processing in the brain.

Although we did not see a three-way interaction, both MDD and cannabis use showed a complete reversal of activity levels relative to their controls in response to the different conditions of the emotion regulation task. Specifically, while participants without MDD showed higher activation to the positive attend condition vs. the other three, those with MDD showed low activation to this condition, with the other three showing higher levels (Figure 3B). Similarly, participants who did not use cannabis showed higher activation levels in response to the negatively vs. positively valenced conditions, while the opposite was true for cannabis users (Figure 3D). The fact that we saw this reversal in all four conditions strongly suggests that both MDD and cannabis use affect several aspects of emotion processing. That is, we observed a change in both positive and negative, and effortful and passive emotion processing. Prior research has shown the effects of MDD and cannabis use on specific types of emotion processing, such as dysfunctional activity during active emotional reappraisal (15, 25). The present results indicate that both MDD and cannabis use may have a more global effect than previously thought.

Both of these effects were observed in the left temporal lobe. While these results were not predicted and are in need of replication, both the left MTG and STG have frequently been associated with emotion processing (31, 71–77), and have previously shown decreases in activity levels in individuals with MDD during emotion processing (25, 77, 78). Both regions are also involved in multisensory association (79). Given that the present stimuli were complex emotional scenes, it is possible that the interactions with MDD and cannabis use in each area reflect differences in multisensory representation. Individuals with MDD showed a reduced representation of positive stimuli during the attend condition, a difference that was eliminated with effortful emotion regulation. Thus, it is possible that individuals with MDD may be successfully augmenting positive representations, while being less successful in their attempt to regulate negative representations. In contrast, cannabis users showed an increased representation of positive stimuli and suppression of negative stimuli, and these mood-altering effects may reflect some of the participants’ motivation for ongoing cannabis use. The difference between the observed effects, namely regulation versus representation of valence, could be why the specific area of temporal lobe differs. Finally, although both MDD and cannabis use affected emotional processing within the temporal lobe, the difference in specific regions may account for why we did not observe a three-way interaction.

Although several regions of the frontal cortex showed activation differences among emotion regulation task conditions, there were no interactions with MDD or cannabis use. Models of both depression and of cannabis use predict the under-activation of frontal regions, specifically the vlPFC, dlPFC, and dmPFC. During healthy emotion regulation, we also observe suppression of these areas (80). Because the individuals with MDD are already experiencing suppression in these regions, it is possible that the amount of change during the emotion regulation task was not enough to appear different from non-depressed participants.

We also found that higher ERQ scores, which represent a greater ability to adaptively control one’s emotions, correlated with less activity in the right frontal lobe. This was observed across all task conditions, indicating that better emotional control leads to less effortful emotion processing overall. While this may seem intuitive, it may be surprising that there was no interaction with condition; for example, Greening and colleagues (25) found suppressed BOLD activity in individuals with MDD during negative regulation compared to healthy controls, but no difference in positive regulation. However, here, even in the ‘attend’ conditions, individuals with low ERQ scores showed more effortful processing than those with high scores. This consistency may reflect that emotion processing occurs even when passively viewing emotionally laden images (81, 82). Poorer emotional regulation has been linked to MDD (83), and correlates with increases in activity in frontal regions when viewing emotional images (80). Thus, these results fit well with previous literature, and suggest that even passive emotional processing is more effortful for those with poorer regulation, which may be a neural representation of less adaptive emotion regulation strategies (77).

The relationship in the left MTG between HAM-D and task condition in individuals with MDD was driven by the steep increase in activity in response to the ‘negative reduce’ condition with increasing score. This relationship echoes the results found when comparing individuals with and without MDD (Fig. 3B), which showed a similar increase in activity in this condition. Notably, a similar relationship was not found in the other three conditions, highlighting the fact that even within a group of persons with MDD, there are individual differences in levels of depressive symptoms that affect different aspects of emotion regulation.

Finally, our emotion regulation task showed activation within the expected network of regions involved in emotion processing, specifically the left inferior parietal lobe, the left middle frontal gyrus, the right insula, and the left inferior frontal gyrus. In both the left inferior parietal lobe and left inferior frontal gyrus, the ‘negative attend’ condition had significantly lower levels of activation than the other conditions. The left middle frontal gyrus showed lower activity to negative versus positive conditions, and the insula showed increased activation in the ‘negative reduce’ condition relative to the others. All four regions have shown differential activation during viewing of emotionally negative stimuli compared to neutral stimuli (71, 84–86), and are thought to belong to a larger network of regions involved in the initial appraisal (inferior frontal gyrus and insula), regulation (middle frontal gyrus), and the final generation of regulated emotional states (inferior parietal lobe) (26, 86). However, although the regions showing an effect of task condition were part of the well-studied emotion processing network, the areas we found to be modulated by MDD, cannabis use, or characteristics of these two factors (e.g., HAM-D score or age of use onset) were outside of this network. The fact that these effects extended beyond typical emotion processing areas during the present task indicates that both MDD and cannabis use have far-reaching consequences for the brain, perhaps affecting domain-general processes (87, 88).

One limitation of the present study is that our analysis of early-onset cannabis use did not identify any significant effects of age of onset, with only a main effect of condition, implying these results were similar across age groups. This was surprising, as early-onset cannabis use was previously associated with increased connectivity between the default mode network and reward-processing areas in the same sample (39), though the early age of onset group was defined differently. Additionally, a recent review paper reported that adolescent exposure to cannabinoids can lead to dysregulation of emotion and reward processing in rats (89). One possible explanation for the lack of effects in this area is the low number of participants in this analysis; only 12 individuals were considered “early” cannabis users, which may not have been a large enough sample to detect differences between early and late cannabis use.

A second limitation is that we did not study the effects of comorbidity with other psychiatric illnesses. Data on comorbidities were collected and reported; as can be seen in Table S1, there was a large range of psychiatric comorbidities within the sample of individuals with MDD. Because of the large variation in the type of comorbidities observed within the sample, we do not have reason to believe that any one diagnosis could be driving the results observed here. However, comorbidity of MDD with other psychopathologies can impact emotion regulation and should be considered in future work.

## 5. Conclusions

As hypothesized, MDD and frequent cannabis use were both associated with functional abnormalities in regions of the brain involved in emotion regulation, but we found that the combination of MDD and cannabis was more complex than strictly additive. We also found that other, related aspects of MDD and cannabis use such as severity of depressive symptoms and emotion regulation style predicted brain activity during emotional processing, highlighting the complex relationship among all three factors. Future studies can continue to examine the role that individual differences have on the relationship between MDD, cannabis use, and the brain.

## Data Availability

Data are available from the corresponding author upon request.

## Acknowledgements

We would like to thank the team at the First Episode Mood and Anxiety Program for providing their hard work with subject recruitment and data collection during the research.

## Author Contributions

RWJN, DGVM, KF, JT, PCW, and EAO conceptualization; MW and ESN, data curation; EAO, Funding acquisition, Supervision; EAO and JT, Methodology; ESN formal analysis; ESN and JP, Writing – original draft; all authors, Writing – review & editing.

## References

1. Kessler RC, et al. (2003) The epidemiology of major depressive disorder. Evidence-Based Eye Care 4(4):186–187.

2. Findlay L (2017) Depression and suicidal ideation among Canadians aged 15 to 24.

3. Behavioral Health Barometer: United States, Volume 5: Indicators as measured through the 2017 National Survey on Drug Use and Health and the National Survey of Substance Abuse Treatment Services. (2019) (Rockville, MD) Available at: https://www.samhsa.gov/data/sites/default/files/Indiana_BHBarometer_Volume_4.pdf.

4. Rush B, et al. (2008) Prevalence of co-occurring substance use and other mental disorders in the Canadian population. Can J Psychiatry 53(12):800–809.

5. Brownlie E, et al. (2019) Early Adolescent Substance Use and Mental Health Problems and Service Utilisation in a School-based Sample. Can J Psychiatry 64(2):116–125.

6. Feingold D, Rehm J, Lev-Ran S (2017) Cannabis use and the course and outcome of major depressive disorder: A population based longitudinal study. Psychiatry Res 251:225–234.

7. Gobbi G, et al. (2019) Association of Cannabis Use in Adolescence and Risk of Depression, Anxiety, and Suicidality in Young Adulthood: A Systematic Review and Meta-analysis. JAMA Psychiatry 76(4):426–434.

8. Henquet C, Krabbendam L, de Graaf R, ten Have M, van Os J (2006) Cannabis use and expression of mania in the general population. J Affect Disord 95(1–3):103–110.

9. Patton GC, et al. (2002) Cannabis use and mental health in young people: cohort study. BMJ 325(7374):1195–1198.

10. van Laar M, van Dorsselaer S, Monshouwer K, de Graaf R (2007) Does cannabis use predict the first incidence of mood and anxiety disorders in the adult population? Addiction 102(8):1251–1260.

11. Wittchen H-U, et al. (2007) Cannabis use and cannabis use disorders and their relationship to mental disorders: a 10-year prospective-longitudinal community study in adolescents. Drug Alcohol Depend 88 Suppl 1:S60–70.

12. Ammerman S, Tau G (2016) Weeding Out the Truth: Adolescents and Cannabis. J Addict Med 10(2):75–82.

13. Lake S, et al. (2020) Does cannabis use modify the effect of post-traumatic stress disorder on severe depression and suicidal ideation? Evidence from a population-based cross-sectional study of Canadians. J Psychopharmacol 34(2):181–188.

14. Schoeler T, et al. (2018) Developmental sensitivity to cannabis use patterns and risk for major depressive disorder in mid-life: Findings from 40 years of follow-up. Psychol Med 48(13):2169–2176.

15. Zimmermann K, et al. (2017) Emotion regulation deficits in regular marijuana users. Hum Brain Mapp 38(8):4270–4279.

16. Cornelis MC, et al. (2019) Age and cognitive decline in the UK Biobank. PLoS One 14(3):1–16.

17. Stephanou K, Davey CG, Kerestes R, Whittle S, Harrison BJ (2017) Hard to look on the bright side: Neural correlates of impaired emotion regulation in depressed youth. Soc Cogn Affect Neurosci 12(7):1138–1148.

18. Dorard G, Berthoz S, Phan O, Corcos M, Bungener C (2008) Affect dysregulation in cannabis abusers: A study in adolescents and young adults. Eur Child Adolesc Psychiatry 17(5):274–282.

19. Gross JJ, John OP (2003) Individual differences in two emotion regulation processes: implications for affect, relationships, and well-being. J Pers Soc Psychol 85(2):348–362.

20. Davidson RJ, Pizzagalli D, Nitschke JB, Putnam K (2002) Depression: perspectives from affective neuroscience. Annu Rev Psychol 53:545–574.

21. Stevens FL, Hurley RA, Taber KH (2011) Anterior cingulate cortex: unique role in cognition and emotion. J Neuropsychiatry Clin Neurosci 23(2):121–125.

22. Mayberg HS, et al. (2005) Deep Brain Stimulation for Treatment-Resistant Depression. Neuron 45(5):651–660.

23. Koenigs M, et al. (2008) Distinct regions of prefrontal cortex mediate resistance and vulnerability to depression. J Neurosci 28(47):12341–12348.

24. Fitzgerald PB, Laird AR, Maller J, Daskalakis ZJ (2008) A meta-analytic study of changes in brain activation in depression. Hum Brain Mapp 29(6):683–695.

25. Greening SG, Osuch EA, Williamson PC, Mitchell DGV (2014) The neural correlates of regulating positive and negative emotions in medication-free major depression. Soc Cogn Affect Neurosci 9(5):628–637.

26. Kohn N, et al. (2014) Neural network of cognitive emotion regulation - An ALE meta-analysis and MACM analysis. Neuroimage 87:345–355.

27. Han T, Alders GL, Greening SG, Neufeld RWJ, Mitchell DGV (2012) Do fearful eyes activate empathy-related brain regions in individuals with callous traits? Soc Cogn Affect Neurosci 7(8):958–968.

28. Ochsner KN, Bunge SA, Gross JJ, Gabrieli JDE (2002) Rethinking feelings: an FMRI study of the cognitive regulation of emotion. J Cogn Neurosci 14(8):1215–1229.

29. Ochsner KN, et al. (2004) For better or for worse: neural systems supporting the cognitive down- and up-regulation of negative emotion. Neuroimage 23(2):483–499.

30. Urry HL, et al. (2006) Amygdala and ventromedial prefrontal cortex are inversely coupled during regulation of negative affect and predict the diurnal pattern of cortisol secretion among older adults. J Neurosci 26(16):4415–4425.

31. Wager TD, Davidson ML, Hughes BL, Lindquist MA, Ochsner KN (2008) Prefrontal-Subcortical Pathways Mediating Successful Emotion Regulation. Neuron 59(6):1037–1050.

32. Brakowski J, et al. (2017) Resting state brain network function in major depression – Depression symptomatology, antidepressant treatment effects, future research. J Psychiatr Res 92:147–159.

33. Kaiser RH, Andrews-Hanna JR, Wager TD, Pizzagalli DA (2015) Large-scale network dysfunction in Major Depressive Disorder: Meta-analysis of resting-state functional connectivity. JAMA Psychiatry 72(6):603–611.

34. Batalla A, et al. (2013) Structural and Functional Imaging Studies in Chronic Cannabis Users: A Systematic Review of Adolescent and Adult Findings. PLoS One 8(2):e55821.

35. Lorenzetti V, et al. (2016) Adolescent Cannabis Use: What is the Evidence for Functional Brain Alteration? Curr Pharm Des 22(42):6353–6365.

36. Wesley MJ, Lile JA, Hanlon CA, Porrino LJ (2016) Abnormal medial prefrontal cortex activity in heavy cannabis users during conscious emotional evaluation. Psychopharmacology (Berl) 233(6):1035–1044.

37. Gruber SA, Rogowska J, Yurgelun-Todd DA (2009) Altered affective response in marijuana smokers: An FMRI study. Drug Alcohol Depend 105(1–2):139–153.

38. Ford KA, et al. (2014) Unique functional abnormalities in youth with combined marijuana use and depression: An fMRI study. Front Psychiatry 5(SEP):1–11.

39. Osuch EA, et al. (2016) Depression, marijuana use and early-onset marijuana use conferred unique effects on neural connectivity and cognition. Acta Psychiatr Scand 134(5):399–409.

40. First MB, Gibbon M, Spitzer RL, Benjamin LS, Williams JBW (1997) Structured Clinical Interview for DSM-IV® Axis II Personality Disorders SCID-II (American Psychiatric Pub).

41. Bava S, Jacobus J, Thayer RE, Tapert SF (2013) Longitudinal changes in white matter integrity among adolescent substance users. Alcohol Clin Exp Res 37 Suppl 1:E181–9.

42. Bolla KI, Brown K, Eldreth D, Tate K, Cadet JL (2002) Dose-related neurocognitive effects of marijuana use. Neurology 59(9):1337–1343.

43. Hamilton M (1960) A rating scale for depression. J Neurol Neurosurg Psychiatry 23:56–62.

44. Youth Risk Behavior Survey (2009) Available at: www.cdc.gov/YRBSS.

45. Lang PJ, Bradley MM, Cuthbert BN (2008) International affective picture system (IAPS): affective ratings of pictures and instruction manual. University of Florida, Gainesville (Tech Rep A-8).

46. Beck AT, Rush AJ, Shaw BF, Emery G (1979) Cognitive therapy of depression (Guilford press, New York).

47. Esteban O, et al. (2020) fMRIPrep: a robust preprocessing pipeline for functional MRI. doi:10.5281/ZENODO.3673912.

48. Esteban O, et al. (2019) fMRIPrep: a robust preprocessing pipeline for functional MRI. Nat Methods 16(1):111–116.

49. Gorgolewski K, et al. (2011) Nipype: A Flexible, Lightweight and Extensible Neuroimaging Data Processing Framework in Python. Front Neuroinform 5:13.

50. Gorgolewski KJ, et al. (2017) Nipype: a flexible, lightweight and extensible neuroimaging data processing framework in Python. 0.13.1. doi:10.5281/ZENODO.581704.

51. Tustison NJ, et al. (2010) N4ITK: Improved N3 bias correction. IEEE Trans Med Imaging 29(6):1310–1320.

52. Avants BB, Epstein CL, Grossman M, Gee JC (2008) Symmetric diffeomorphic image registration with cross-correlation: Evaluating automated labeling of elderly and neurodegenerative brain. Med Image Anal 12(1):26–41.

53. Reuter M, Rosas HD, Fischl B (2010) Highly accurate inverse consistent registration: a robust approach. Neuroimage 53(4):1181–1196.

54. Dale AM, Fischl B, Sereno MI (1999) Cortical surface-based analysis. I. Segmentation and surface reconstruction. Neuroimage 9(2):179–194.

55. Klein A, et al. (2017) Mindboggling morphometry of human brains. PLoS Comput Biol 13(2):e1005350.

56. Fonov V, Evans A, McKinstry R, Almli C, Collins D (2009) Unbiased nonlinear average age-appropriate brain templates from birth to adulthood. Neuroimage 47:S102.

57. Zhang Y, Brady M, Smith S (2001) Segmentation of brain MR images through a hidden Markov random field model and the expectation-maximization algorithm. IEEE Trans Med Imaging 20(1):45–57.

58. Greve DN, Fischl B (2009) Accurate and robust brain image alignment using boundary-based registration. Neuroimage 48(1):63–72.

59. Jenkinson M, Bannister P, Brady M, Smith S (2002) Improved Optimization for the Robust and Accurate Linear Registration and Motion Correction of Brain Images. Neuroimage 17(2):825–841.

60. Cox RW, Hyde JS (1997) Software tools for analysis and visualization of fMRI data. NMR Biomed 10(4-5):171–178.

61. Power JD, Schlaggar BL, Petersen SE (2014) Studying Brain Organization via Spontaneous fMRI Signal. Neuron 84(4):681–696.

62. Behzadi Y, Restom K, Liau J, Liu TT (2007) A component based noise correction method (CompCor) for BOLD and perfusion based fMRI. Neuroimage 37(1):90–101.

63. Lanczos C (1964) Evaluation of Noisy Data. J Soc Ind Appl Math Ser B Numer Anal 1(1):76–85.

64. Abraham A, et al. (2014) Machine learning for neuroimaging with scikit-learn Front Neuroinformatics 8:14.

65. Cox RW (1996) AFNI: Software for Analysis and Visualization of Functional Magnetic Resonance Neuroimages. Comput Biomed Res 29(3):162–173.

66. Gold S, et al. (1998) Functional MRI statistical software packages: a comparative analysis. Hum Brain Mapp 6(2):73–84.

67. Chen G, Saad ZS, Britton JC, Pine DS, Cox RW (2013) Linear mixed-effects modeling approach to FMRI group analysis. Neuroimage 73:176–190.

68. Bagiella E, Sloan RP, Heitjan DF (2000) Mixed-effects models in psychophysiology. Psychophysiology 37(1):13–20.

69. Baayen RH, Davidson DJ, Bates DM (2008) Mixed-effects modeling with crossed random effects for subjects and items. J Mem Lang 59(4):390–412.

70. Tibon R, Levy DA (2015) Striking a balance: analyzing unbalanced event-related potential data. Front Psychol (JAN):1–4.

71. Goldin PR, McRae K, Ramel W, Gross JJ (2008) The Neural Bases of Emotion Regulation: Reappraisal and Suppression of Negative Emotion. Biol Psychiatry 63(6):577–586.

72. Sato W, Kochiyama T, Yoshikawa S, Naito E, Matsumura M (2004) Enhanced neural activity in response to dynamic facial expressions of emotion: An fMRI study. Cogn Brain Res 20(1):81–91.

73. Chan SWY, Norbury R, Goodwin GM, Harmer CJ (2009) Risk for depression and neural responses to fearful facial expressions of emotion. Br J Psychiatry 194(2):139–145.

74. Cancelliere AEB, Kertesz A (1990) Lesion localization in acquired deficits of emotional expression and comprehension. Brain Cogn 13(2):133–147.

75. Jenkins LM, et al. (2017) Integrated cross-network connectivity of amygdala, insula, and subgenual cingulate associated with facial emotion perception in healthy controls and remitted major depressive disorder. Cogn Affect Behav Neurosci 17(6):1242–1254.

76. Sakaki M, Niki K, Mather M (2012) Beyond arousal and valence: the importance of the biological versus social relevance of emotional stimuli. Cogn Affect Behav Neurosci 12(1):115–139.

77. Pico-Perez M, Radua J, Steward T, Menchon JM, Soriano-Mas C (2017) Emotion regulation in mood and anxiety disorders: A meta-analysis of fMRI cognitive reappraisal studies. Prog Neuropsychopharmacol Biol Psychiatry 79(Pt B):96–104.

78. Keedwell PA, Andrew C, Williams SCR, Brammer MJ, Phillips ML (2005) The neural correlates of anhedonia in major depressive disorder. Biol Psychiatry 58(11):843–853.

79. Cappe C, Rouiller EM, Barone P (2009) Multisensory anatomical pathways. Hear Res 258(1–2):28–36.

80. Abler B, et al. (2010) Habitual emotion regulation strategies and depressive symptoms in healthy subjects predict fMRI brain activation patterns related to major depression. Psychiatry Res-Neuroimaging 183(2):105–113.

81. Hall LMJ, et al. (2014) An FMRI study of emotional face processing in adolescent major depression. J Affect Disord 168:44–50.

82. Pine DS, et al. (2001) Cortical brain regions engaged by masked emotional faces in adolescents and adults: An fMRI study. Emotion 1(2):137–147.

83. Sakakibara R, Kitahara M (2016) The relationship between Cognitive Emotion Regulation Questionnaire (CERQ) and depression, anxiety: Meta-analysis. Shinrigaku Kenkyu 87(2):179–17985.

84. Van Dillen LF, Heslenfeld DJ, Koole SL (2009) Tuning down the emotional brain: An fMRI study of the effects of cognitive load on the processing of affective images. Neuroimage 45(4):1212–1219.

85. Grecucci A, Giorgetta C, Bonini N, Sanfey AG (2013) Reappraising social emotions: The role of inferior frontal gyrus, temporo-parietal junction and insula in interpersonal emotion regulation. Front Hum Neurosci 7(SEP):1–12.

86. Grecucci A, Giorgetta C, Van’t Wout M, Bonini N, Sanfey AG (2013) Reappraising the ultimatum: An fMRI study of emotion regulation and decision making. Cereb Cortex 23(2):399–410.

87. Neta M, Kelley WM, Whalen PJ (2013) Neural Responses to Ambiguity Involve Domain-general and Domain-specific Emotion Processing Systems. J Cogn Neurosci 25(4):547–557.

88. Waters AC, Mayberg HS (2017) Brain-based biomarkers for the treatment of depression: Evolution of an idea. J Int Neuropsychol Soc 23(9-10 Special Issue):870–880.

89. Renard J, Rushlow WJ, Laviolette SR (2016) What can rats tell us about adolescent cannabis exposure? Insights from. Can J Psychiatry 61(6):328–334.

