## Supplemental materials for "Emotion regulation in emerging adults with major depressive disorder and frequent cannabis use"

| **Table S1.** Current comorbidities within the sample,  determined using the Structured Clinical Interview for Diagnosis, Clinical Version | |
| --- | --- |
| **Comorbidity** | **n (%)** |
| Social Phobia | 9 (12.3%) |
| ^a^Drug Abuse/Dependence | 9 (12.3%) |
| Alcohol Abuse/Dependence | 8 (11.0%) |
| PTSD | 6 (8.2%) |
| Generalized Anxiety Disorder | 4 (5.5%) |
| Specific Phobia | 2 (2.7%) |
| Binge Eating Disorder | 1 (1.4%) |
| Body Dysmorphia | 1 (1.4%) |
| Hypochondriasis | 1 (1.4%) |
| Panic Attacks | 1 (1.4%) |
| Panic Disorder with Agoraphobia | 1 (1.4%) |
| Somatization | 1 (1.4%) |
| ^a^Drugs other than cannabis | |
